# External Validation Of A Novel Signature Of Illness In Continuous Cardiorespiratory Monitoring To Detect Early Respiratory Deterioration Of ICU Patients

**DOI:** 10.1101/2021.05.24.21257711

**Authors:** Rachael A. Callcut, Yuan Xu, J Randall Moorman, Christina Tsai, Andrea Villaroman, Anamaria J. Robles, Douglas E Lake, Xiao Hu, Matthew T Clark

**Affiliations:** University of California, Davis, Department of Surgery, Davis, CA, USA; University of California, San Francisco, Department of Surgery, San Francisco, CA, USA; University of Virginia, UVa Center for Advanced Medical Analytics, Charlottesville, VA, USA; University of Virginia, Cardiovascular Division, Charlottesville, VA, USA; Duke University, School of Nursing; Advanced Medical Predictive Devices, Diagnostics, and Displays, Charlottesville, VA

**Author notes:** **CORRESPONDING AUTHOR: Name:** J Randall Moorman, MD, **Affiliation:** University of Virginia, **Address:** Box 800158, Charlottesville VA 22947, **Email:**. **EMAIL ADDRESSES Rachael A Callcut:**. **Yuan Xu:**. **Christina Tsai:**. **Andrea Villaroman:**. **Anamaria Robles:**. **Doug Lake:**. **Matthew Clark:**. **J Randall Moorman:**. **Xiao Hu:**.

## Abstract

The goal of predictive analytics monitoring is the early detection of patients at high risk of subacute potentially catastrophic illnesses. An excellent example of a targeted illness is respiratory failure leading to urgent unplanned intubation, where early detection might lead to interventions that improve patient outcomes. Previously, we identified signatures of this illness in the continuous cardiorespiratory monitoring data of Intensive Care Unit patients and devised algorithms to identify patients at rising risk. Here, we externally validated three logistic regression models to estimate the risk of emergency intubation developed in Medical and Surgical ICUs at the University of Virginia. We calculated the model outputs for more than 8000 patients in the University of California – San Francisco ICUs, 240 of whom underwent emergency intubation as determined by individual chart review. We found that the AUC of the models exceeded 0.75 in this external population, and that the risk rose appreciably over the 12 hours before the event. We conclude that there are generalizable physiological signatures of impending respiratory failure in the continuous cardiorespiratory monitoring data.

## INTRODUCTION

Patients in the intensive care unit (ICU) that experience respiratory failure leading to emergent intubation have significantly longer hospital length-of-stay and higher in-hospital mortality. ^1–3^ Earlier identification of patients that will require intubation may allow earlier intervention to reduce morbidity and mortality. Patients in the early stages of respiratory failure might be given corticosteroids, bronchodilators, or supplemental oxygen, or placed on non-invasive positive-pressure ventilation. ^4^ Patients for whom non-invasive positive-pressure ventilation is insufficient, or recently extubated patients that require re-intubation, might be intubated electively rather than emergently. ^5,6^ Timely intervention aimed at better preparation for intubation, team coordination, proper intubation medication selection, and avoidance of peri-intubation hypotension improve outcomes of patients requiring emergent intubation. These observations have never been more true than they are now, during the COVID-19 pandemic when emergency intubation endangers the providers as well as the patients. ^7^

Predictive analytics monitoring strives to identify high-risk patients earlier by providing continuous risk estimates to clinical personnel in real-time. These early warning signals can allow intervention to alter the patient’s trajectory in a more favorable direction. In a prospective study using analogous predictive analytics for sepsis that were displayed at the bedside of preterm infants, mortality was reduced by more than 20%. ^8,9^ Similarly, in adults, predictive analytics for sepsis may reduce rates of septic shock and associated mortality by up to 50%. ^10,11^

Candidate risk marker models for subacute potentially catastrophic illnesses like respiratory failure leading to urgent unplanned intubation ^3,12^ are usually based only on static demographics and comorbidities, and intermittent vital signs and laboratory tests. ^13^ As a result, these models may reflect the decisions of clinicians more so than changes in patient physiology. ^14^ That is, if the Electronic Health Record (EHR) shows that a clinician ordered a stat blood gas and chest X-ray, is it really a prediction to say that respiratory failure is imminent? If the physician thought of it first, do these analytics really represent the leading indicators of a patient’s illness, or are they just the lagging indicators of clinicians’ actions? On the other hand, models based on continuous cardiorespiratory dynamics from bedside physiological monitors have the advantage of reporting on the patient’s condition directly. Clinician-initiated interventions do not directly influence them.

Before using a predictive analytical model for prospective clinical practice, it is vital to validate that model externally across different patient populations and institutions. ^15,16^ Different care units and institutions may have substantially different distributions of demographics, socio-economic groups, admitting practices, and care patterns, all of which may degrade the calibration and discriminatory performance of a model. This study tested the hypothesis that predictive dynamic analytical models for respiratory failure leading to urgent unplanned intubation using continuously available data from ubiquitous physiologic monitors are well-suited for application at an external center.

## METHODS

### Study Design

We retrospectively studied a cohort of patients admitted to an ICU at the University of California San Francisco Medical Center (UCSF). We studied admissions to the two mixed medical/surgical ICUs, two neurological ICUs (NICUs), and coronary care (CCU) ICUs. Each ICU has continuous physiological monitoring archived by BedMaster (Hillrom, Chicago IL). The primary outcome was respiratory failure leading to urgent unplanned intubation.

### Study population and primary outcome

We studied consecutive ICU admissions from May 1, 2013, through April 30, 2015, and selected patients intubated in the ICU. We excluded intubation events before ICU admission (*e*.*g*., in the operating room or emergency department) and patients with “Do Not Resuscitate” or “Do Not Intubate” orders (DNR/DNI). We classified events as planned (elective) or unplanned (emergent). Planned intubations included those done for procedures (such as endoscopy, interventional radiology procedures, or preceding elective operations) and others documented to be elective. We considered all others to be unplanned. We examined the procedure notes and the physician notes in individual medical records to determine the reason for intubation. We extracted the timing of intubation from the nursing and respiratory therapist’s documentation. Two independent practitioners independently reviewed each potential case of emergent intubation.

### Identification of mechanically ventilated patients

We excluded data for patients during epochs when they were already mechanically ventilated. Thus, it was necessary to know the time of all intubations and extubations (not only the emergent intubations). While we knew the times of emergent intubations, all other intubation and all extubation times were not available. We instead used ventilator respiratory rate flowsheet entries from respiratory therapists (RT) as a surrogate to identify periods of mechanical ventilation, merging the results with known times of emergent intubations. We reasoned that this parameter, which specifies that the respiratory rate was measured from the ventilator, was a sufficiently reliable indicator of the presence of mechanical ventilation.

We extracted the total ventilator respiratory rate flowsheet entry documented by RTs. Figure 1(left) shows the probability density of time between RT documentation: most RT documentation of the ventilator respiratory rate occurred more frequently than every 6 hours, consistent with clinical practice and affirming the utility of defining mechanical ventilation this way. Mechanical ventilation was defined as starting at the first recording of ventilator respiratory rate and ending at the last ventilator respiratory rate; see Figure 1 (right) example patient 1. We split the period of mechanical ventilation when the time between consecutive measurements from a patient was larger than 16 hours and identified the patient as not ventilated in the interim; see Figure 1 (right) example patient 2. Isolated measurements (*i*.*e*., those without another measurement within 16 hours) were used to define the start of a ventilation epoch with a duration of 1 hour; see Figure 1(right) example patient 3. For emergently intubated patients, we verified that the epochs of mechanical ventilation started at the time of intubation. When emergent intubation occurred in the middle of an automatically determined ventilation epoch, we split the mechanical ventilation epoch defining the time of extubation as the time of the preceding ventilator respiratory rate and the time of intubation as the time of emergent intubation.

**Figure 1:**
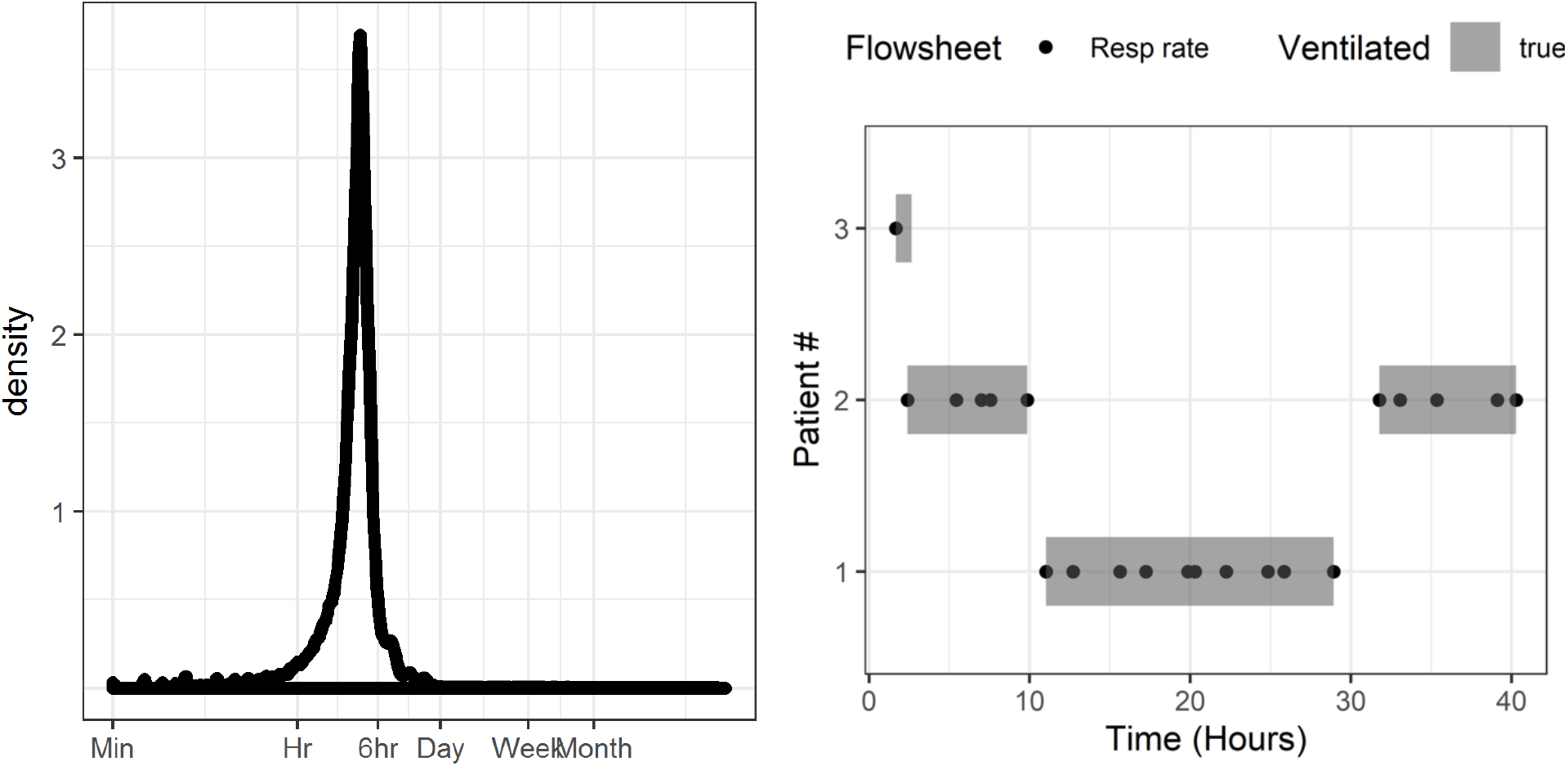
(left) Probability density of the time between consecutive ventilator respiratory rate entries for all patients. (right) Simplified examples showing the aggregation of flowsheet ventilator respiratory rate (points) into epochs of mechanical ventilation (shaded rectangles). All measurements within 6 hours are combined into a single epoch (patient 1), whereas measurements separated by more than 16 hours are split into multiple epochs (patient 2) and isolated measurements are defined as 1-hour epochs (patient 3)

### Predictors of emergent intubation

We calculated three risk estimates for emergent intubation. ^1,2^ The models were developed on data from SICU and MICU patients at the University of Virginia (UVa) Health System and use only continuous cardiorespiratory dynamics calculated from the bedside physiological monitors. From Politano *et al*. ^2^ we used the vital signs only model. This model was developed on a subset of surgical ICU patients, excluding those on ancillary services. We also used the SICU and MICU models of Moss *et al*. ^1^ We previously validated the model of Politano at UVa for predicting upgrade from surgical intermediate care unit (IMU) to ICU, with and without intubation. ^17^ The SICU model of Moss was also validated at the same institution as part of a model for identifying low-risk patients at the time of surgical IMU and ICU discharge. ^18^

All three models are binary logistic regression. The Politano model uses linear relationships between predictors and the response, while the Moss models include cubic splines to allow for non-linear relationships. The Politano model was built with a forward stepwise procedure, while the Moss models use a pre-specified feature set based on prior clinical knowledge. The output of each model is the estimated probability of emergent intubation, though with different time horizons: 24 hours for the Politano model, 4 hours for the Moss MICU model, and 6 hours for the Moss SICU model.

Model inputs are were calculated in 30-minute windows with 50% overlap: means and standard deviations of, and cross-correlations between, vital signs (heart rate, respiratory rate, peripheral oxygen saturation, and blood pressure), as well as statistical measures of cardiac dynamics (slope of log RR interval variance versus log scale for detrended fluctuation analysis, ^19^ coefficient of sample entropy, ^20^ and the standard deviation of RR intervals).

Missing data were imputed with the median values from the UVa development cohort. We divided the estimated probability by the average probability of emergent intubation in the training set to yield the fold-increase in the probability of the event, which we label relative risk. Features and models were calculated using CoMET® (AMP3D, Charlottesville, VA). No features or models were available to the care team, and all patients received standard of care. The UCSF Institutional Review Board approved this study with a waiver of consent.

### Statistical analysis

We evaluated the performance of the UVa urgent intubation risk models for identifying UCSF patients before emergent intubation. We used the continuous risk estimates from the three models and, where appropriate, a binary response variable. The response was defined as “1” for event patients during the time window immediately preceding the emergent intubation (the event population). The response was defined as “0” for those patients that were never emergently intubated (control patients) as well as for event patients far from the time of emergent intubation.

We analyzed the dynamic changes of each model near the time of emergent intubation. We aligned the every-15-minute risk estimates relative to the time of emergent intubation (defined as time zero). For each model, at each 15-minute epoch, we calculated the average predicted risk for all event patients with data and plotted the average time series relative to the time of emergent intubation. We used the Wilcoxon sign rank test every 15 minutes to test the null hypothesis that risk estimates were equal to risk estimates for the same patient 12 hours prior. At time points where we rejected the null hypothesis at 0.05 significance level, we interpreted this as a significant change in estimated risk that may have provided an early warning.

We evaluated the discriminatory value of each model using the area under the receiver operating characteristic (AUC), also known as the C-statistic. We evaluated the performance of risk estimates for varying event time window definitions: window start times ranged from 4 to 24-hours before emergent intubation. The window ended at the time of emergent intubation. Confidence intervals were determined by 200 bootstrap runs, resampled by hospital admission.

Finally, we evaluated the calibration of each model. For each model, we calculated the deciles of predicted relative risk. We calculated the observed risk in each decile as the proportion of measurements that corresponded to times within 12 hours of emergent intubation and divided by the average risk from the training set. We then plotted the observed vs. predicted relative risk: perfectly calibrated models fall on the line of identity.

## RESULTS

We studied 9,828 admissions for 8,434 patients to UCSF. There were 240 episodes of emergent intubation in 238 hospital admissions. There was continuous monitoring data before 225 (93%) of events. Table 1 shows the characteristics of the study population. Patients with emergent intubation were less likely to be white, more likely to be Asian, had 3-fold higher mortality, and stayed 19 days longer in hospital. We calculated 105.5 patient-years of risk estimates: the three risk estimates were calculated for 3.7 million 15-minute epochs. We censored 32.3% of measurements for patients who were already mechanically ventilated or patients following DNI orders. The incidence of emergent intubation was 0.9 per 100 ICU days, significantly lower than in the Moss ^1^ and Politano ^2^ development cohorts (2.1 and 2.8 per 100 ICU days, respectively).

**Table 1:**
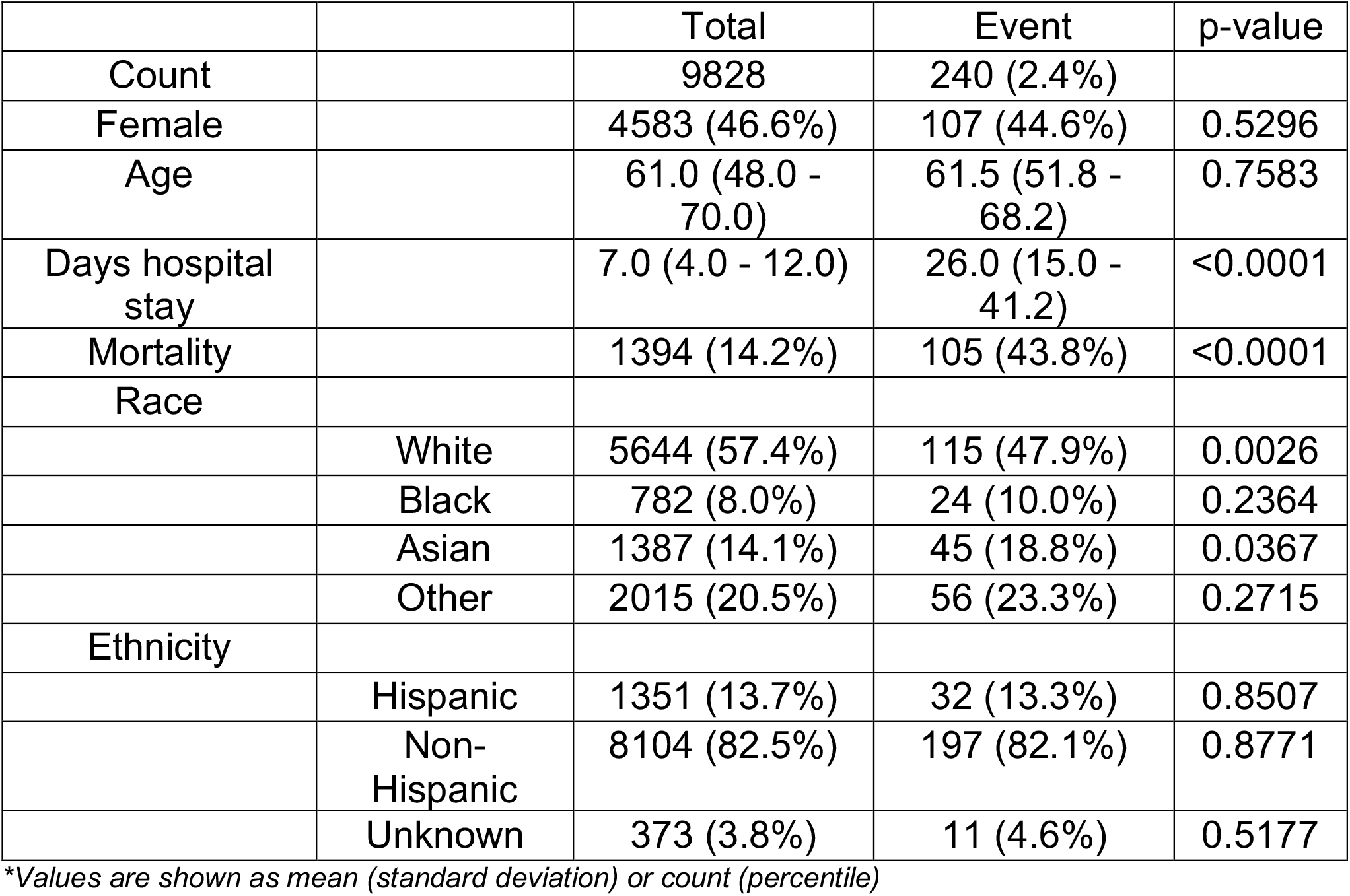
Characteristics of the study population

Figure 2 shows the length of time patients were in the ICU and not mechanically ventilated before emergent intubation: about 120 patients were in the ICU and not mechanically ventilated at least 24 hours before the event. The Figure also shows the number of events with continuous monitoring data available for modeling. Figure 3 shows the time course of risk estimates for UCSF patients using the three UVa models for urgent unplanned intubation during the 48 hours preceding emergent intubation. Risk estimates doubled over the 12 to 24 hours before emergent intubation, from about 1.5 to more than 3. At each time, we performed a signed-rank test with the null hypothesis that risk estimates are equal to risk estimates from the same patient 12 hours prior. White points identify times when we rejected the null hypothesis. Risk estimates were significantly higher beginning about 5 hours before emergent intubation.

**Figure 2:**
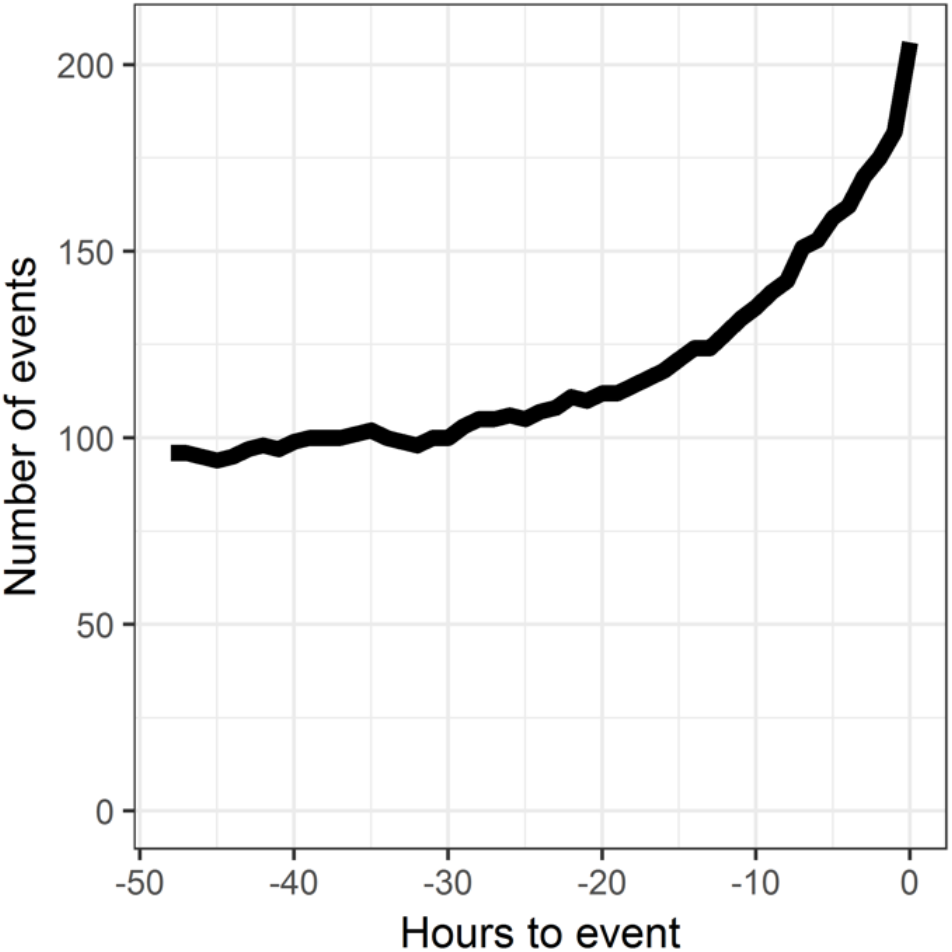
Number of events with continuous monitoring data as a function of time leading up to emergent intubation.

**Figure 3:**
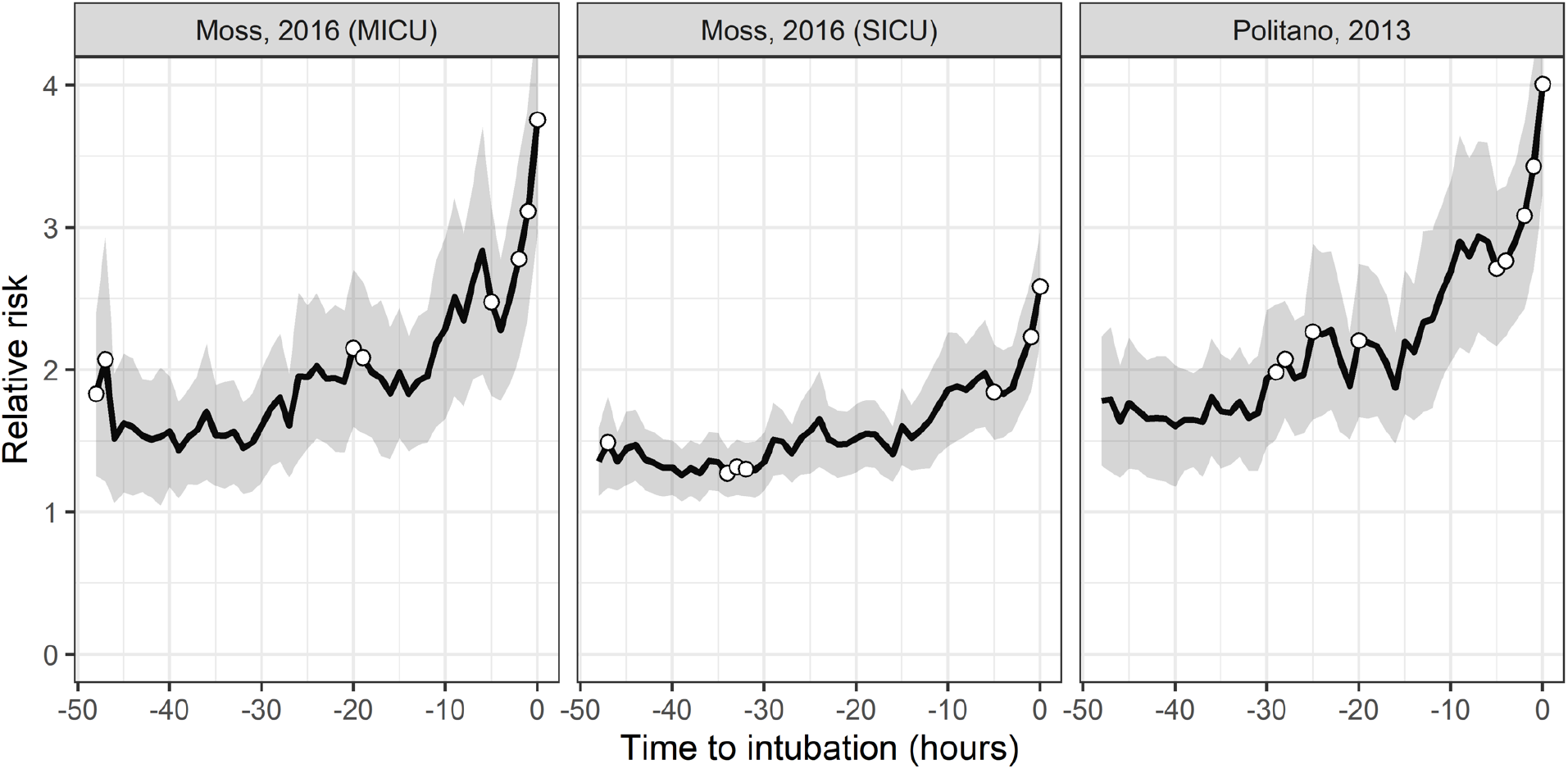
Average time course of risk estimates over the 48 hours leading up to the time of emergent intubation. Relative risk is the fold-increase in the probability of an event with respect to average. The gray ribbon is the 95% confidence interval around the mean. White points indicate that the risk estimates at that time are significantly higher (p < 0.05) than risk estimates 12 hours prior.

Figure 4 shows the performance of the three models as a function of the time window before emergent intubation. All the data from patients without emergent intubation served as ‘control.’ Thus, for a 24-hour detection window, the data for event patients within 24 hours of emergent intubation were identified as ‘event,’ and the data from event patients far from the event were ‘control.’ The AUC rose from about 0.75 to about 0.78 for event windows 24 to 4 hours, respectively. The MICU model from Moss had slightly better performance than the SICU models from Moss or Politano.

**Figure 4:**
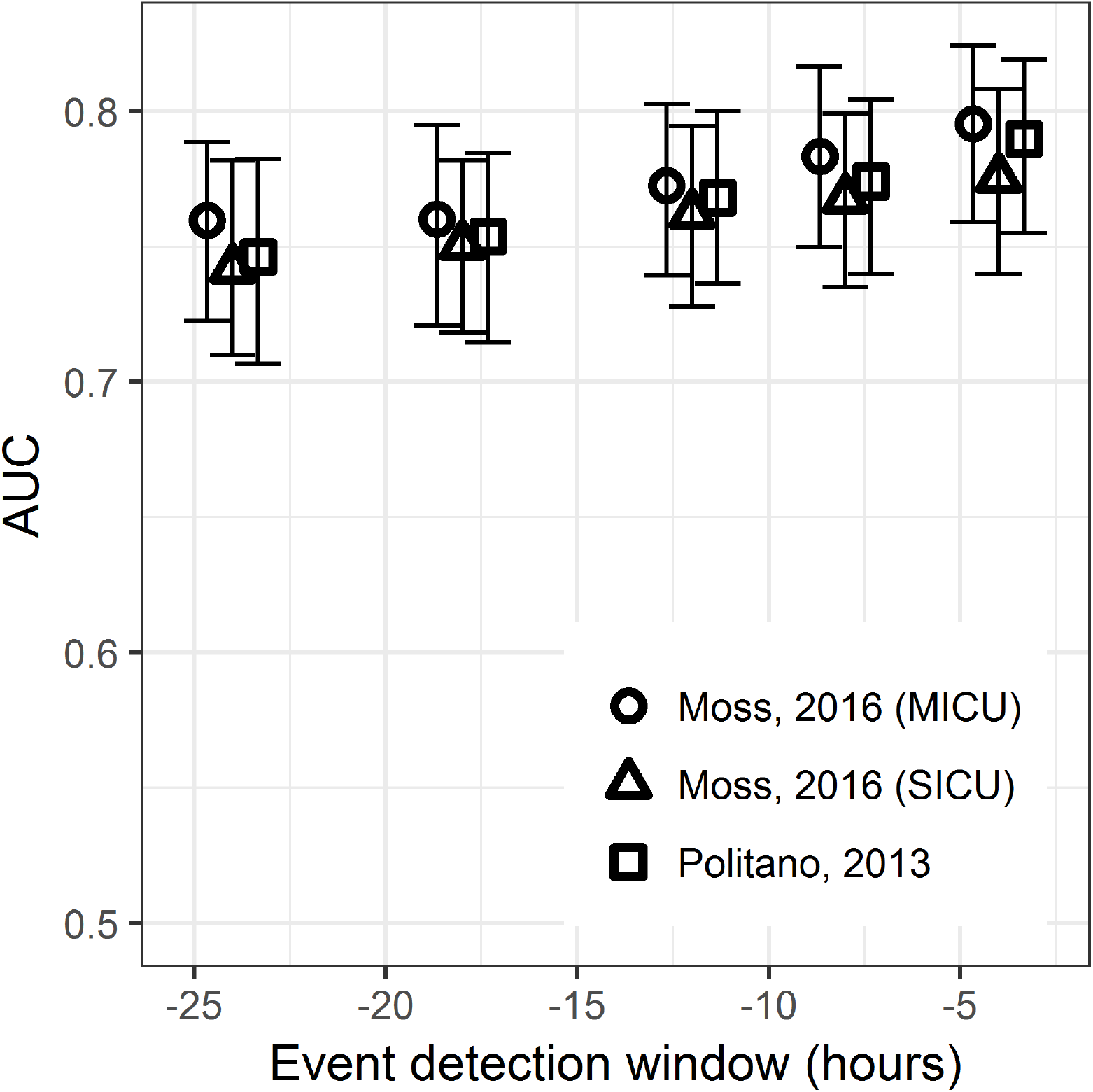
Area under the receiver operating characteristic as a function of the window size before emergent intubation defined as the event, from 4 to 24 hours. The 95% confidence interval is indicated by error bars and was determined by 200 bootstrap runs resampled by admission.

Figure 5 shows the calibration of the three models using a 12-hour detection window. Well-calibrated models have predicted risk equal to observed risk (dashed line). The SICU models from Moss and Politano exhibited excellent calibration in all but the highest risk patients, where they overestimated the risk. The MICU intubation model from Moss also overestimated risk for high-risk patients and deviated from the line of identity in the two lowest deciles of predicted risk.

**Figure 5:**
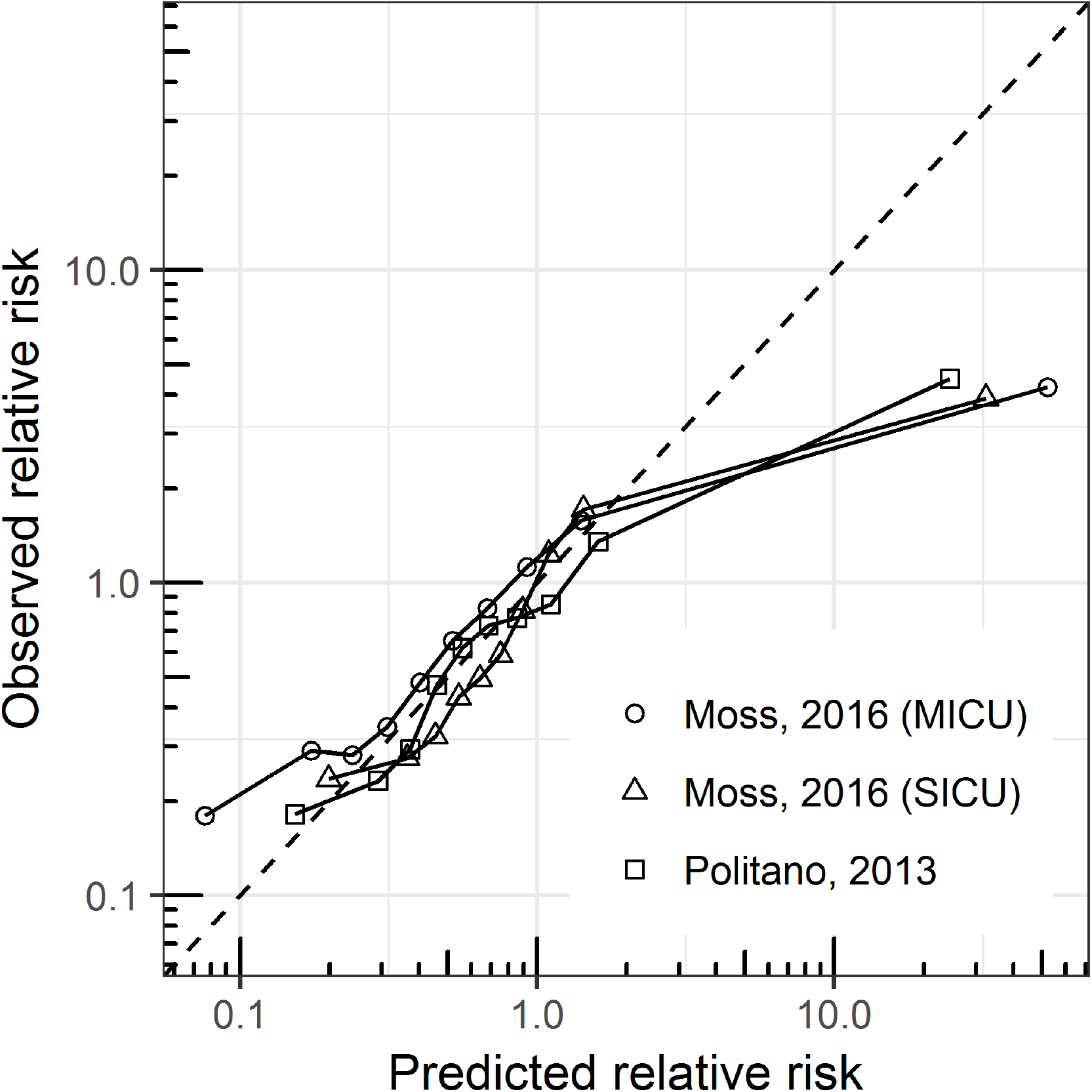
Calibration curves for the three models for emergent intubation. The observed relative risk is plotted as a function of the predicted risk. Each point represents 10% of the data, and the line of identity (perfect calibration) is shown as a dashed line.

## DISCUSSION

This study evaluated the performance of 3 predictive analytical models for early detection of respiratory failure leading to emergent intubation at an external center. These models are based solely upon analyses of continuous cardiorespiratory monitoring data to detect physiologic signatures of illness. The existing models were initially developed to predict emergent intubation in the UVa SICU (two models) and MICU (one model). This study sought to externally validate the performance using an independent cohort of patients from all ICUs University of California San Francisco. The major findings were that the models accurately identified patients at risk for emergent intubation (AUC > 0.75 starting at 12 hours pre-intubation), risk estimates continued to rise until the time of emergent intubation, and these predictive models were well-calibrated.

Whereas elective intubation in the operating room is a highly safe procedure, emergent intubation outside the operating room is commonly associated with complications. ^21–23^ These complications include hemodynamic compromise or severe hypoxemia leading to cardiac arrest, esophageal intubation, aspiration, and pneumothorax. ^22,24^ Cardiac arrest attributable to intubation complications occurs in 2-4% of emergent intubations, and few of these patients survive to hospital discharge even when initially resuscitated. ^23–25^ Identifying patients earlier who are at risk of needing emergent intubation may be vital in avoiding these poor outcomes. ^23,26^

Recent studies link poor outcomes to potentially mitigatable factors that are time-dependent. These include a worse outcome for patients intubated around the change of nursing shift. ^22^ Wardi *et al*. proposed that this finding resulted from potential staff fatigue, hand-off errors, and lack of familiarity with a patient, which led to overlooking subtle but important changes in patient condition. ^22^ For each of these potential explanations, predictive algorithms such as those validated in this current study might overcome these challenges at vulnerable moments for our critically ill patients. Essentially, these algorithms convert complex data that is difficult for a clinician to assimilate in real-time into risk estimates based on detecting illness-specific signatures. The result is an early warning signal displayed at the bedside that can draw attention to critical changes in the patient’s condition. Here, we found that the pathophysiological signature of respiratory failure in medical and surgical ICU training sets transferred well to identify emergent intubation in the medical, surgical, neurological, and coronary care ICU cohorts of the validation set. This comports with the findings of Moss *et al*., who found that respiratory failure had a similar signature of illness between surgical and medical ICU patients. ^1^

Additionally, our data affirm that the clinical deterioration before respiratory failure leading to urgent unplanned intubation is often a slowly progressive process over many hours. This time frame creates an opportunity to perform interventions that would make emergent intubation safer and even possibly avoided. A pre-intubation checklist integrating interventions to treat hemodynamic instability led to fewer complications. ^22,23,27^ This is consistent with investigations that have correlated the worse outcomes in emergent intubation in the Emergency Department (ED) occurring in those that develop hemodynamic compromise following intubation. ^24,28^

Commonly, patients requiring emergent intubation are in shock, and an increased shock index highly correlates with intubation-associated cardiac arrest. ^22,28^ Volume depletion, vasodilation, acidosis, and reduced venous return resulting from increased positive pressure ventilation are all factors that contribute. Each of these could be addressed, albeit to differing degrees of success, in the lead time provided by an early warning signal. Trivedi *et al*. argue that the shock index (the ratio of maximum heart rate to lowest systolic blood pressure) is a helpful adjunct to intervene in the immediate 60 minutes preceding intubation. ^28^ However, they acknowledge that addressing the “dynamic changes in patient status in the ICU… require continuous monitoring and interpretation of data before the development of overt hypotension and cardiorespiratory collapse.”

We propose that implementation and integration of predictive analytics monitoring into clinical practice may provide an opportunity for timely clinical action, as the prediction of respiratory failure leading to urgent unplanned intubation can be present and growing for hours pre-intubation. We cannot definitively determine from this study if interventions during this window would avoid intubation, but it would likely make them safer. Both of these possibilities warrant further investigation in a prospective study. Any such prospective study should leverage processes for optimal integration of predictive analytics for evoking clinical action. ^26^

This study was limited in several ways. We relied on surrogate data to initially identify patients who had mechanical ventilation initiated during their ICU stay. It is possible that due to documentation errors, the location of intubation initiation could have been misclassified. However, to avoid this, each potential case of ICU intubation was reviewed by two clinicians to verify the location of intubation, the reason for intubation, and the timing of intubation. Similarly, we attempted to identify tracheostomy patients that may go on and off mechanical ventilation and excluded those with mechanical ventilation initiation following tracheostomy.

Our results show that all models are well-calibrated for the lowest 90% of predicted risk but overestimate the risk in the highest decile: patients in the highest decile have 3-to 4-fold higher risk of respiratory failure than average, but the estimated risk is 30-to 40-fold higher than average. Accurate calibration at low and moderate risks may allow these models to be used for accurate clinical assessment of patient status as well as response to interventions. Overestimation at higher risk may limit such uses at these levels but still may draw attention to high-risk patients as intended. For practical purposes, it may be useful to cap predicted risk estimates for clinical implementation to mitigate this issue.

Determining the exact timing of emergent intubation was challenging in this study. Physician documentation through intubation procedure notes was not a reliable source of the timing as it was apparent in chart review that this documentation is often delayed. This is understandable as the physician’s attention around the event is focused on providing bedside care. To address this, we confirmed the time of intubation with the first nursing documentation of administered intubation-related medications cross-referenced with the respiratory therapist documentation of initial ventilator settings. In all cases, these times were within 5 minutes of each other. A more accurate evaluation of the lead time of the prediction would be to use the exact moment that the clinical decision was made to intubate. We do not know this, of course, but we know from the clinical review of the records that the actual intubation came only a short time later. We note that any delay makes the predictive model look better because the worsening derangements of the patient status make for higher risk estimates. We defend the practice as the best available and superior to a standard method of using the highest model estimate observed during the hospitalization, even if the model estimate took place weeks before the event.

In addition, we did not quantify the performance of predictive models in the context of known risk factors: diagnoses, demographics, or severity of illness. ^3,12^ We note, however, that these known risk factors are static indicators; though they may identify high-risk patients, they do not rise leading up to the time of lung failure. Finally, we did not evaluate other predictors of emergent intubation, such as shock index from vital signs and laboratory measurements. Adding independent streams of information improves performance, ^29^ though we found that plug-and-play models with bedside physiological monitors have excellent performance.

## CONCLUSION

Earlier identification of signatures of illness using continuous cardiorespiratory monitoring that arise from subtle changes in physiologic deterioration may provide a useful adjunct to clinicians to mitigate need for emergent intubation.

## Data Availability

N/A

## AUTHOR CONTRIBUTIONS

Study Design: Callcut, Clark, Moorman, Hu

Data Analysis: Callcut, Xu, Clark, Lake, Moorman, Hu

Data Interpretation: Callcut, Xu, Tsai, Villaroman, Robles, Lake, Clark, Moorman, Hu Writing: Callcut, Clark

Critical Review: Callcut, Xu, Tsai, Villaroman, Robles, Lake, Clark, Moorman, Hu

## Notes

**CONFLICTS OF INTEREST:** Moorman and Clark are officers and own stock in Advanced Medical Predictive Devices, Diagnostics, and Displays Inc

**FINANCIAL DISCLOSURES:** Dr. Callcut is supported by a Career Development Award from the NIH Big Data to Knowledge Initiative (NIH K01ES026834).

### Competing Interest Statement

Moorman and Clark are officers and own stock in Advanced Medical Predictive Devices, Diagnostics, and Displays, Charlottesville, VA.

### Funding Statement

No extramural funding

### Author Declarations

Approved by IRBs at UCSF and the University of Virginia with waiver of consent

### Summary of Updates

Minor text revisions; addition of a second panel to Figure 1; change in author order.

